## Supplementary Information for "Epydemix: An open-source Python package for epidemic modeling with integrated approximate Bayesian calibration"

#### Contents

**S1 Stochastic SIR Model: Example** **2**

**S2 ABC Algorithms** **4**

**S3 Modeling COVID-19 in Massachusetts** **6**

S3.1 Epidemic model . . . . . . . . . . 6

S3.2 Model calibration . . . . . . . . . . 8

### S1 Stochastic SIR Model: Example

As example for the Epydemix simulation paradigm, we consider the prototypical SIR model. Susceptible and healthy individuals are placed in the $S$ compartment. Through interactions with infected individuals ($I$), they can become infected with probability $\beta$. After the infectious period ($\mu^{-1}$), infected individuals transition to the recovered compartment ($R$), where they can no longer spread the disease or become reinfected. This model can be translated into the following stochastic processes, which can be iterated computationally:

$$\begin{aligned} S_k(t+\delta t) &= S_k(t) - Bin(S_k(t), \lambda_r(k,t)), \\ I_k(t+\delta t) &= I_k(t) - Bin(I_k(t), \mu_r), \\ R_k(t+\delta t) &= R_k(t) + Bin(I_k(t), \mu_r), \end{aligned} \tag{1}$$

where $\lambda_r(k,t)$ and $\mu_r$ are the transition probabilities obtained by transforming rates into risks. Specifically, $\mu_r = 1 - e^{-\mu\delta t}$ and $\lambda_r(k,t) = 1 - e^{-\lambda(k,t)\delta t}$, where $\lambda(k,t)$ represents the force of infection for group $k$, i.e., the per-capita rate at which susceptible individuals in group $k$ transition to the infected compartment. In Epydemix, forces of infection (i.e., transition rates of mediated transitions) are computed by accounting for interactions with individuals in mediating compartment through a contact matrix $\mathbf{C}$. The element $C_{kk'}$ of the matrix describes the average number of contacts an individual in group $k$ has with individuals in group $k'$ per unit of time (set here to 1 day). In this context, $\lambda(k,t)$ is calculated as:

$$\lambda(k,t) = \beta \sum_{k'=1}^{K} C_{kk'} \frac{I_{k'}(t)}{N_{k'}}, \tag{2}$$

where $N_{k'}$ is the total number of individuals in group $k'$. In Epydemix, the simulation of this process is achieved by iteratively solving the equations in System 1, calculating the appropriate transition probabilities for binomial sampling at each step. Algorithm S1 provides the pseudocode for the simulation of this system.

**Algorithm S1** Stochastic SIR Model Simulation

**Input:**
  $S_k(0)$, $I_k(0)$, $R_k(0)$: Initial compartment values for all groups $k$
  $C_{kk'}$: Contact matrix
  $\beta$: Transmission rate
  $\mu$: Recovery rate
  $\delta t$: Time step
  $T$: Total simulation time
**Output:** $S_k(t)$, $I_k(t)$, $R_k(t)$ for all $t$
Initialize $S_k(0)$, $I_k(0)$, $R_k(0)$ for all groups $k$
Set $t \leftarrow 0$
**while** $t < T$ **do**
  **for** each group $k$ **do**
    Compute force of infection: $\lambda(k,t) \leftarrow \beta \sum_{k'=1}^{K} C_{kk'} \frac{I_{k'}(t)}{N_{k'}}$
    Compute infection probability: $\lambda_r(k,t) \leftarrow 1 - e^{-\lambda(k,t)\delta t}$
    Sample new infections: new_infections $\leftarrow \text{Bin}(S_k(t), \lambda_r(k,t))$
    Compute recovery probability: $\mu_r \leftarrow 1 - e^{-\mu \delta t}$
    Sample recoveries: new_recoveries $\leftarrow \text{Bin}(I_k(t), \mu_r)$
    Update compartments:

$$\begin{aligned} S_k(t+\delta t) &\leftarrow S_k(t) - \text{new\_infections} \\ I_k(t+\delta t) &\leftarrow I_k(t) + \text{new\_infections} - \text{new\_recoveries} \\ R_k(t+\delta t) &\leftarrow R_k(t) + \text{new\_recoveries} \end{aligned}$$

  **end for**
  Increment time: $t \leftarrow t + \delta t$
**end while**
**return** $S_k(t)$, $I_k(t)$, and $R_k(t)$ for all $t$

### S2 ABC Algorithms

In this section we provide pseudocodes for the three ABC algorithms: ABC Rejection Algorithm (Algorithm S2), ABC Algorithm with Simulation Budget (Algorithm S3), ABC-SMC Algorithm (Algorithm S4).

**Algorithm S2** ABC Rejection Algorithm

**Input:**
  $P$: Population size (number of accepted parameter sets)
  $\pi(\boldsymbol{\theta})$: Prior distribution of parameters
  $d(\cdot, \cdot)$: Distance metric
  $\epsilon$: Tolerance threshold
  $f(\cdot|\boldsymbol{\theta})$: Simulator model
  $\mathbf{y}_{\text{obs}}$: Observed data
**Output:** Set of accepted parameters $\{\boldsymbol{\theta}_i\}_{i=1}^{P}$
Initialize an empty set of accepted parameters: Accepted $\leftarrow \{\}$
**while** $|\text{Accepted}| < P$ **do**
  Sample a parameter set $\boldsymbol{\theta}_i$ from the prior distribution $\pi(\boldsymbol{\theta})$
  Simulate $\mathbf{y}_i \sim f(\cdot|\boldsymbol{\theta}_i)$
  Compute the distance $d(\mathbf{y}_i, \mathbf{y}_{\text{obs}})$
  **if** $d(\mathbf{y}_i, \mathbf{y}_{\text{obs}}) < \epsilon$ **then**
    Add $\boldsymbol{\theta}_i$ to Accepted
  **end if**
**end while**
**return** Accepted

**Algorithm S3** ABC Algorithm with Simulation Budget

**Input:**
  $B$: Total simulation budget (number of total simulations allowed)
  $X$: Percentage of best trajectories to select
  $\pi(\boldsymbol{\theta})$: Prior distribution of parameters
  $d(\cdot, \cdot)$: Distance metric
  $f(\cdot|\boldsymbol{\theta})$: Simulator model
  $\mathbf{y}_{\text{obs}}$: Observed data
**Output:** Set of accepted parameters $\{\boldsymbol{\theta}_i\}_{i=1}^{\lfloor B \cdot X/100 \rfloor}$
Initialize an empty set of parameter-distance pairs: Candidates $\leftarrow \{\}$
**for** $i = 1$ to $B$ **do**
  Sample a parameter set $\boldsymbol{\theta}_i$ from the prior distribution $\pi(\boldsymbol{\theta})$
  Simulate $\mathbf{y}_i \sim f(\cdot|\boldsymbol{\theta}_i)$
  Compute the distance $d(\mathbf{y}_i, \mathbf{y}_{\text{obs}})$
  Add pair $(\boldsymbol{\theta}_i, d(\mathbf{y}_i, \mathbf{y}_{\text{obs}}))$ to Candidates
**end for**
Sort Candidates by the distance $d(\mathbf{y}_i, \mathbf{y}_{\text{obs}})$ in ascending order
Select the top $\lfloor B \cdot X/100 \rfloor$ parameter sets based on the sorted distances
**return** Selected parameter sets $\{\boldsymbol{\theta}_i\}_{i=1}^{\lfloor B \cdot X/100 \rfloor}$

**Algorithm S4** ABC-SMC Algorithm

**Input:**
  $P$: Number of particles
  $\delta$: Sequence of tolerance levels $\delta_1 > \delta_2 > \cdots > \delta_T$
  $\pi(\boldsymbol{\theta})$: Prior distribution
  $f(\cdot|\boldsymbol{\theta})$: Simulator model
  $d(\cdot,\cdot)$: Distance function
  $K(\cdot|\cdot)$: Perturbation kernel
  $\mathbf{y}_{\text{obs}}$: Observed data
**Output:** Posterior distribution approximations $\{\boldsymbol{\theta}_i^{(t)}\}_{i=1}^{P}$ for each $t = 1, \ldots, T$
**for** $t = 1$ to $T$ **do**
  **if** $t = 1$ **then**
    **for** $i = 1$ to $P$ **do**
      **repeat**
        Sample $\boldsymbol{\theta}_i^{(1)} \sim \pi(\boldsymbol{\theta})$
        Simulate $\mathbf{y}_i \sim f(\cdot|\boldsymbol{\theta}_i^{(1)})$
      **until** $d(\mathbf{y}_i, \mathbf{y}_{\text{obs}}) \leq \epsilon_1$
      Set weight $w_i^{(1)} = \frac{1}{P}$
    **end for**
  **else**
    **for** $i = 1$ to $P$ **do**
      **repeat**
        Sample $\boldsymbol{\theta}_i^{(t-1)} \sim \{\boldsymbol{\theta}_j^{(t-1)}\}_{j=1}^{N}$ with weights $w_j^{(t-1)}$
        Perturb $\boldsymbol{\theta}_i^{(t)} \sim K(\boldsymbol{\theta}|\boldsymbol{\theta}_i^{(t-1)})$
        Simulate $\mathbf{y}_i \sim f(\cdot|\boldsymbol{\theta}_i^{(t)})$
      **until** $d(\mathbf{y}_i, \mathbf{y}_{\text{obs}}) \leq \epsilon_t$
      Compute weight $w_i^{(t)} = \frac{\pi(\boldsymbol{\theta}_i^{(t)})}{\sum_{j=1}^{N} w_j^{(t-1)} K(\boldsymbol{\theta}_i^{(t)}|\boldsymbol{\theta}_j^{(t-1)})}$
    **end for**
  **end if**
  Normalize weights: $w_i^{(t)} \leftarrow \frac{w_i^{(t)}}{\sum_{j=1}^{N} w_j^{(t)}}$ for $i = 1, \ldots, P$
**end for**

### S3 Modeling COVID-19 in Massachusetts

#### S3.1 Epidemic model

In this section, we provide a detailed description of the epidemic model introduced in the main text, which is used to simulate the spread of COVID-19 in Massachusetts, US, during early 2020.

The natural history of the disease is represented using an SEIR-like compartmental model, augmented with additional compartments to account for disease-related deaths. We stratify the population into 10 age groups $k$ (i.e., $[0-9, 10-19, 20-24, 25-29, 30-39, 40-49, 50-59, 60-69, 70-79, 80+]$), where each group is characterized by a population size $N_k$, reflecting the real demographic distribution. Additionally, we incorporate the contact matrix $\mathbf{C} \in \mathbb{R}^{K \times K}$, where the element $C_{i,j}$ represents the average number of daily effective contacts between individuals in age group $i$ and those in age group $j$ [1].

The transition rate of susceptible individuals $S$ to the exposed state $E$, known as the force of infection, is given by:

$$\lambda(k,t) = \beta s(t) r(t) \sum_{k'=1}^{K} C_{kk'} \frac{I_{k'}(t)}{N_{k'}}. \tag{3}$$

Here, the infection rate is assumed to be proportional to the fraction of infectious individuals in each age group, where $\beta$ denotes transmissibility of the disease. The expression also includes a seasonality modulation term $s(t)$, which accounts for variations in humidity, temperature, and other environmental factors that influence both transmissibility and contact patterns [2, 3]. The seasonality function is defined as:

$$s_i(t) = \frac{1}{2}\left[\left(1 - \frac{s_{min}}{s_{max}}\right) \sin\left(\frac{2\pi}{365}(t - t_{max,i}) + \frac{\pi}{2}\right) + 1 + \frac{s_{min}}{s_{max}}\right] \tag{4}$$

where $i$ represents the Hemisphere, and $t_{max,i}$ corresponds to the time of maximum seasonality. We set this to January $15^{th}$ in the Northern Hemisphere and shift it by six months in the Southern Hemisphere, assuming no seasonality in tropical regions. The maximum seasonality factor is fixed at $s_{max} = 1$, while $s_{min}$ is treated as a free parameter [3, 4].

The force of infection is further modulated by the contact variation factor $r(t)$, which is derived from the Community Mobility Report published by Google LLC [5]. This dataset provides percentage changes in mobility and visits to various locations over time. Since our model does not distinguish between different locations (i.e., contexts), we introduce a single mobility parameter, $m(t)$, representing the average percentage reduction in visits across all locations (excluding parks due to their atypical behavior). The mobility factor is converted into an effective contact reduction parameter as:

$$r(t) = \left(1 - \frac{|m(t)|}{100}\right)^2. \tag{5}$$

The rationale behind this formulation is that under a homogeneous mixing assumption, the number of potential contacts $C$ scales quadratically with population size, i.e., $C = N(N-1)/2 \sim N^2$. If the number of individuals present in a location is reduced to $N'(t) = (1 - m(t)/100)N$, the number of potential contacts follows $C'(t) \sim N'(t)^2 = (1 - m(t)/100)^2 N^2$, yielding the contact reduction factor $r(t) = C'(t)/C$.

Exposed individuals progress to the infectious state $I$ at rate $\epsilon$, which is the inverse of the latent period. Infectious individuals then transition to the removed state $R$ at rate $\mu$, the inverse of the infectious period. The removed compartment includes individuals who are no longer infectious.

To model COVID-19 mortality, we track the transitions from $I_k$ to $R_k$. A fraction of these transitions, determined by age-stratified estimates of the Infection Fatality Rate (IFR) from Ref. [6], leads to the $D_{0,k}$ compartment, which represents deceased individuals. Since death does not occur immediately after recovery due to hospitalization, isolation, and reporting delays, we introduce additional compartments $(D_{1,k}, D_{2,k}, D_{3,k}, D_{4,k})$. Individuals transition between these compartments at a rate of $4/\Delta$, where $\Delta$ is the average delay (in days) between recovery and recorded death. The number of individuals transitioning from $D_{3,k}$ to $D_{4,k}$ on day $t$ corresponds to the reported deaths on that day. The introduction of multiple $D_{i,k}$ compartments ensures that the delay distribution follows an Erlang distribution with shape parameter $n = 4$ [7].

The disease progression is simulated using stochastic chain binomial processes, as described above. The model can be expressed by the following stochastic process that can be conveniently iterated computationally:

$$\begin{aligned}
S_k(t+\delta t) &= S_k(t) - Bin(S_k(t), \lambda_r(k,t)), && (6)\\
E_k(t+\delta t) &= E_k(t) + Bin(S_k(t), \lambda_r(k,t)) - Bin(E_k(t), \epsilon_r), && (7)\\
I_k(t+\delta t) &= I_k(t) + Bin(E_k(t), \epsilon_r) - Mult_1(I_k(t), 1 - e^{-\mu(1-IFR_k)\delta t}, 1 - e^{-\mu IFR_k \delta t})\\
&\quad - Mult_2(I_k(t), 1 - e^{-\mu(1-IFR_k)\delta t}, 1 - e^{-\mu IFR_k \delta t}),\\
R_k(t+\delta t) &= R_k(t) + Mult_1(I_k(t), 1 - e^{-\mu(1-IFR_k)\delta t}, 1 - e^{-\mu IFR_k \delta t}),\\
D_{0,k}(t+\delta t) &= D_{0,k}(t) + Mult_2(I_k(t), 1 - e^{-\mu(1-IFR_k)\delta t}, 1 - e^{-\mu IFR_k \delta t}) - Bin(D_{0,k}(t), 4/\Delta),\\
D_{1,k}(t+\delta t) &= D_{1,k}(t) + Bin(D_{0,k}(t), 4/\Delta) - Bin(D_{1,k}(t), 4/\Delta),\\
D_{2,k}(t+\delta t) &= D_{2,k}(t) + Bin(D_{1,k}(t), 4/\Delta) - Bin(D_{2,k}(t), 4/\Delta),\\
D_{3,k}(t+\delta t) &= D_{3,k}(t) + Bin(D_{2,k}(t), 4/\Delta) - Bin(D_{3,k}(t), 4/\Delta),\\
D_{4,k}(t+\delta t) &= D_{4,k}(t) + Bin(D_{3,k}(t), 4/\Delta)
\end{aligned}$$

For the above contagion process, the basic reproduction number is $R_0 = \rho(\tilde{\mathbf{C}})\beta/\mu$, where $\tilde{C_{ij}} = C_{ij}N_i/N_j$, $\rho(\cdot)$ is the spectral radius [8], and the generation time of the disease is $T_G = \epsilon^{-1} + \mu^{-1}$.

##### S3.2 Model calibration

The model is calibrated to weekly reported deaths from Ref. [9] using the ABC-SMC algorithm, employing 10 generations with $1,000$ accepted particles per generation. The free parameters and their prior distributions, all assumed to follow uniform distributions, are: the basic reproduction number $R_0 \sim U(1.5, 6.0)$, the seasonality parameter $s(t) \sim U(0.6, 1.0)$, the delay between recovery and death $\Delta \sim U(7, 35)$, and the initial number of infected individuals across age groups $I_0 \sim U(10, 10000)$.

Figure S1 presents the estimated posterior distributions for these parameters.

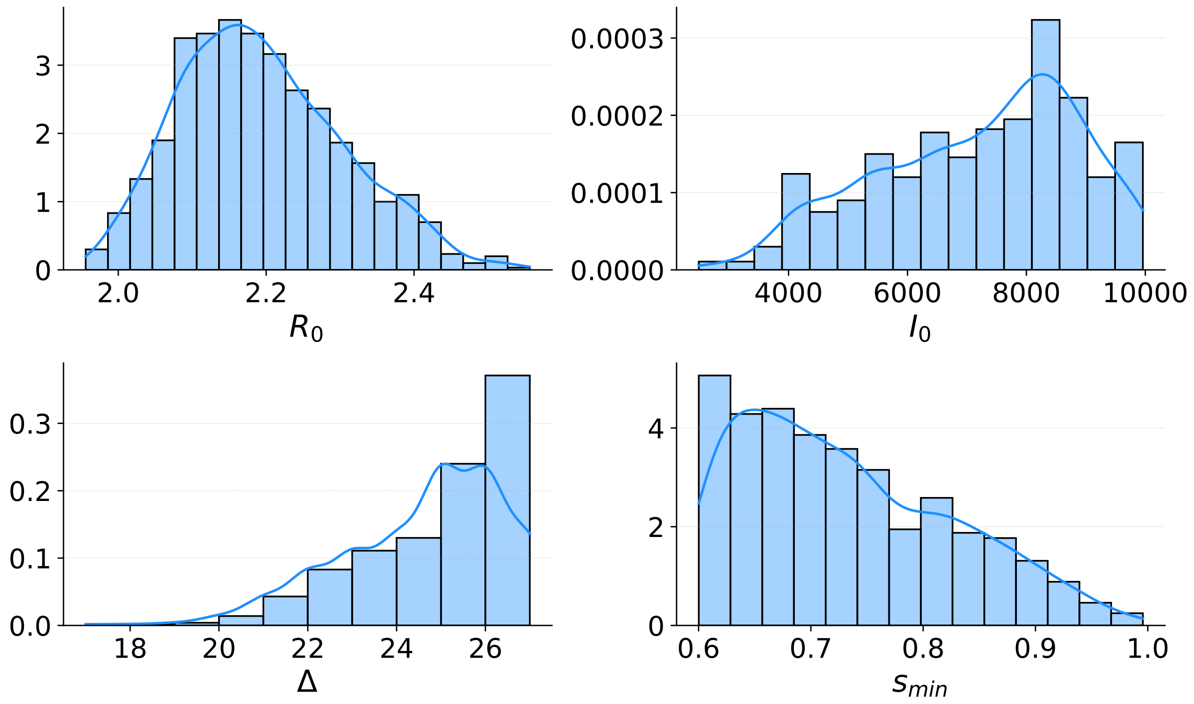


Figure S1: Posterior distributions of free epidemiological parameters inferred via ABC-SMC calibration. The histograms represent the sampled posterior distributions for the basic reproduction number ($R_0$), initial number of infected individuals ($I_0$), average delay between recovery and death ($\Delta$), and minimum seasonality factor ($s_{min}$). The blue curves indicate kernel density estimates (KDE) of the posterior distributions.
